# Impact of a Pharmacy Care Management Service on Health Spending, Resource Use, and Medication Adherence for Medically Complex Medicare Advantage Members

**DOI:** 10.1101/2023.04.12.23287952

**Authors:** Loren Lidsky, Shawn Hallinan, Josh Benner, Stephen Jones, Elise Smith, Patricia Houck, Heidi Stevenson, Don Yoder, Chronis Manolis, Allen Naidoo, Niteesh Choudhry, Chester B Good

**Author notes:** CORRESPONDING AUTHOR Loren Lidsky, MS.

## Abstract

**BACKGROUND:** The Medicare Advantage (MA) program has grown rapidly over the past two decades, particularly among medically complex beneficiaries, which has implications for healthcare utilization, costs, and quality in the MA program. Ensuring safe and effective medication use in this population has been identified as a priority by policymakers, yet there is limited evidence to guide MA plans’ pharmacy care management efforts.

**OBJECTIVE:** To evaluate the impact of an integrated pharmacy care management (PCM) program implemented at a regional Medicare Advantage and Part D (MAPD) plan on 12-month cost outcomes, health services use, and medication adherence quality measures for polypharmacy members enrolled in the program as well as for a subgroup predicted to have high potential cost savings from improved medication adherence.

**METHODS:** We conducted a retrospective cohort study using adjudicated administrative claims data and multi-stage matching methods. The PCM program was offered by telephone to MAPD members who had filled eight or more chronic medications in the 180 days prior to being screened for eligibility. The PCM cohort consisted of individuals enrolled in the PCM program who filled at least one prescription after enrollment, had no evidence of hospice care, and were continuously eligible for health plan benefits for at least 12 months before and 12 months after enrollment. Potential controls were members who met the same criteria but who did not participate in the PCM program and filled prescriptions at non-PCM pharmacies. A commercially available algorithm (the Value of Future Adherence [VFA] score) was used to predict potential future cost savings associated with improved medication adherence for all members at baseline. Control members were matched to PCM members in a 5:1 ratio using a two-stage matching process. Outcomes were measured over 12 months and included per enrollee per month (PEPM) health spending, health resource utilization, and medication adherence for oral diabetes medications, renin-angiotensin system antagonists, and statins. Outcomes were assessed for all members and in the subgroup with high VFA scores.

**RESULTS:** A total of 724 PCM members matched to 3,620 control members, with 196 members in the high VFA subgroup. Among all PCM members, there was a $50 (95% CI: $15, $86; p=0.005) PEPM increase in average pharmacy spending compared to controls, and an offsetting $158 (95% CI: -$265, -$51; p=0.004) PEPM decrease in average medical spending, resulting in a $108 (95% CI: -$221, $5; p=0.062) PEPM lower average total cost of care in PCM members after 12 months. Savings were driven primarily by the high VFA subgroup, which incurred an average of $52 (95% CI: -$19, $130; p=0.187) PEPM greater pharmacy spending and $458 (95% CI: -$678, -$233; p<0.001) PEPM less medical spending than controls, for an average decrease of $406 (95% CI: -$645, -$161; p<0.001) PEPM in total cost of care. PCM members experienced a 15% (p=0.008) reduction in inpatient stays compared to control members; high VFA PCM members had 32% (p<0.001) fewer inpatient stays. The PCM program was associated with increases in the percent of members adherent to renin-angiotensin system antagonist, statin, and oral antidiabetic therapy, ranging from 7.3 to 12.9 (p<0.001) percentage points among all PCM members and 12.2 to 14.3 (p<0.05) percentage points for high VFA members.

**CONCLUSION:** The PCM program was associated with significantly lower medical spending, reduced hospital admissions, and improved adherence to chronic medications in members receiving the program for 12 months. Benefits of the program are greatest among members in the high VFA subgroup. Our findings support the value of an integrated pharmacy care management program in the polychronic MAPD population and underscore the value of targeting the PCM program to members predicted to benefit the most rather than merely on the basis of the number of medications a member is taking.

## BACKGROUND

The Medicare Advantage (MA) program has grown rapidly over the past two decades. Managed Medicare plans provided coverage to 28 million members in 2022 (45% of all Medicare beneficiaries) and are expected to enroll more beneficiaries than traditional fee-for-service Medicare in 2023.^1^ Major contributors to the recent growth are medically complex beneficiaries who need institutional-level care, have severe or disabling chronic conditions, or are dually eligible for Medicare and Medicaid.^1^

The shift toward more medically complex beneficiaries has implications for healthcare utilization, costs, and quality in the MA program. Average total spending per MA member grew by 13% from 2012 to 2015, with key drivers being increased spending on prescription drugs (38% higher), hospital stays (21% higher), and skilled nursing facilities (20% higher).^2^ MA plans’ performance on many Star Ratings quality measures improved from 2012 to 2015, but measures of adherence to blood pressure, cholesterol, and diabetes medications declined. This has prompted calls for MA plans to develop targeted interventions to prevent medical complications and to reduce spending on post-acute care by increasing the quality of medication use.^2^

Many population health management programs used by MA plans include services aimed at improving medication prescribing and adherence, such as medication reconciliation, use of clinical pharmacists to educate members about their regimens, and provider alerts about medication adherence.^3^ However, these services often are one-time encounters delivered after hospitalization or non-adherence already has occurred and are independent of beneficiaries’ usual pharmacy services. As a result, while such interventions may reduce inappropriate prescribing or increase adherence over the short-term, evidence of meaningful effects on hospital admissions and health care costs has been conflicting or of low quality, suggesting a need for more sustained and integrated interventions, better targeting, and more rigorous outcomes studies.^4-7^

We designed, implemented, and evaluated an integrated pharmacy care management (PCM) program for MA members with multiple chronic conditions and elevated risk of avoidable and costly health services. The PCM program combined multiple interventions suggested in previous studies to improve health outcomes through more effective prescribing or adherence: medication reconciliation,^8-9^ refill synchronization,^5^ adherence packaging,^10^ home delivery,^11^ monthly medication reviews with member education,^12^ and coordination among prescribing healthcare providers.^13^ We implemented the PCM program at a regional Medicare Advantage and Part D (MAPD) plan. This study reports 12-month cost outcomes, health services use, and medication adherence quality measures for all PCM members compared to a matched control group of members who received usual pharmacy care. To assess the value of targeting the PCM program, a commercially available algorithm (the Value of Future Adherence [VFA] score)^14-16^ was used to predict potential future cost savings associated with improved medication adherence for all members at baseline. Outcomes were measured separately for the subgroup of PCM members with high VFA scores and their matched controls.

## METHODS

### Setting and Design

We conducted a retrospective cohort study utilizing multi-stage matching methods to evaluate the PCM program’s effect on health services utilization and total cost of care for medically complex members of UPMC Health Plan’s MAPD plans. This study was a program evaluation by UPMC Health Plan and thus deemed exempt by the UPMC Institutional Review Board.

### Subject Eligibility and Intervention

The PCM program was offered to members of UPMC MAPD plans who had filled eight or more chronic medications in the 180 days prior to being screened for eligibility. Eligibility rules for the PCM program excluded individuals enrolled in a state-sponsored pharmaceutical assistance program or using a pharmacy known to provide services similar to the PCM program. Members were screened for eligibility and offered the PCM program by telephone on a rolling basis starting in December 2019; the last date of enrollment for members included in this evaluation was November 2021.

Members who enrolled in the PCM program participated in a telephonic onboarding appointment to obtain a medication history, reconcile current prescriptions, and synchronize medication refills. To provide comprehensive and consistent pharmacy care for enrolled members, a single pharmacy dispensed medications and provided clinical pharmacy services for the PCM program. Members were contacted monthly to assess changes to health status and the medication regimen. As-needed clinical pharmacy services included member and caregiver education, referral to health care providers, consultation with prescribers on changes to the medication regimen, and referral to social support or case management programs provided by the MA plan. The PCM program also included specialized dispensing services and home delivery in monthly cycles to minimize waste. Medications were sorted into pouches corresponding to the day and time of each dose. A large label with photographs and instructions for each medication was affixed to the box containing the monthly supply of pouches.

### Study Cohorts

The PCM cohort consisted of individuals who agreed to enroll in the PCM program, had no evidence of hospice care, filled at least one prescription after enrollment, remained enrolled for at least 12 months, and were continuously eligible for health plan benefits for at least 12 months before and 12 months after enrollment in the PCM program. Potential controls were beneficiaries who were eligible for the PCM program but could not be reached or were reached and declined to enroll, and subsequently filled a medication at a non-PCM pharmacy.

Control members were required to meet the same criteria as PCM members. The follow-up period began on the index date, which was the date of the first prescription fill after enrollment (PCM members) or the first prescription fill after outreach was attempted (control members).

### Baseline Covariates

We measured baseline characteristics in the 12-month period prior to the index date for PCM and control members. These included age, gender, index date, receipt of low-income subsidy, number of chronic medications, Charlson Comorbidity Index score, health services use (emergency department visits and hospitalizations), total inpatient days, baseline pharmacy spending, baseline medical spending, and the proportion of members who completed a comprehensive medical review (CMR) in the past 12 months. PCM and control members were scored at baseline on the VFA score, a commercially available algorithm that predicts potential cost savings from improving medication adherence.^14-16^ Medical and pharmacy cost data were winsorized to the 99th percentile to reduce distortion due to outliers.

To reduce further the potential for selection bias in our results, we employed a two-stage matching process to ensure control members were similar to PCM members on variables known to be associated with health care spending and PCM enrollment. First, PCM members and all potential controls were directly matched on age, pre-index hospitalization, pre-index total medical costs and total pharmacy costs, level of VFA score, and CMR completion. Next, we estimated each PCM and control member’s propensity to enroll in the PCM program based on these same six variables used in the direct match along with the remaining covariates and matched five control members to each PCM member using a greedy matching approach, with calipers of width equal to 0.2 of the standard deviation of the logit of the propensity score.^17,18^ Standardized mean differences (SMD) were used to compare the PCM and control groups after matching, with an SMD of <10% considered well-matched.^19,20^ The c-statistic from a post-match propensity model served as another indication of the effectiveness of the match, with a value approaching 0.5 suggesting that the measured characteristics of a PCM member and a control member are indistinguishable.^21^

### Outcomes

We used adjudicated claims data to evaluate cost, healthcare utilization, and medication adherence outcomes during the 12-month follow-up period. Per enrollee per month (PEPM) health spending was measured using total allowed amounts (plan plus member paid amounts) for all pharmacy and medical utilization in the 12-month follow-up period; pharmacy spending was normalized to the plan’s preferred retail rates for all members. Health services use was estimated by measuring inpatient stays, emergency department visits, and urgent care visits during the follow-up period. Medication adherence was assessed for oral diabetes medications, renin-angiotensin system antagonists, and statins by calculating the proportion of days covered (PDC) using CMS Medicare Star Ratings measure specifications.^22^ Accordingly, to be eligible for the adherence evaluation, members had to have filled two or more prescriptions in the drug class at any point during the 12-month period prior to the baseline period. Members were considered adherent if their PDC for a given class was ≥80% during the follow-up period. All outcomes were compared between the overall PCM members and their matched controls and separately for the subgroup of PCM members with high VFA scores and their matched controls.

### Statistical Analyses

All outcomes were measured at baseline and in the follow-up period within PCM and control groups. A difference-in-difference calculation was performed to adjust for residual baseline variation between the PCM and control cohorts. The results from this final calculation were reported as the absolute difference in the tables. To provide scope and scale for utilization differences, we additionally reported a relative difference (as a percentage). This was defined as the ratio of the absolute difference to baseline PCM utilization, adjusted for 12-month pre-post changes in control utilization. Since PCM and control members were not matched on baseline adherence rates, differences in adherence between groups were estimated using a binomial generalized linear model with identity link and robust standard errors that adjusted for the interaction between time (pre / post index) and cohort. Due to the skewness of cost and utilization data, confidence intervals and p-values were estimated using bootstrapping (10,000 iterations) with replacement. All analyses were conducted using SAS^®^ version 9.4 software (SAS, Cary, NC).

## RESULTS

There were 744 PCM and 15,411 potential control members prior to matching, of whom 207 and 3,684 were in the high VFA PCM and control subgroups, respectively; 70% of the PCM cohort who met health plan eligibility also met the 12-month PCM program continuous enrollment requirement **(Appendix Figure e1)**. Baseline characteristics of PCM members and potential controls prior to matching are shown in the Appendix **(Table e2)**. Compared to eligible members who did not enroll in the PCM program, the PCM members tended to be younger, female, and to receive low-income subsidy. PCM members also were more likely than potential controls to have had a recent ED visit and a comprehensive medical review.

Matching criteria were satisfied for 724 (97%) of PCM members and 196 (95%) of the high VFA subgroup. PCM and control members were well-matched with respect to demographic characteristics, baseline health services use and costs of care, and previous engagement in comprehensive medication reviews, with all SMD values <10% and c-statistics of 0.527 (all members) and 0.570 (high VFA members) **(Table 1)**.

**Table 1.** Baseline characteristics of Pharmacy Care Management members and controls, after matching.

| Baseline Characteristics | All Members |  |  | High VFA Members |  |  |
| --- | --- | --- | --- | --- | --- | --- |
|  | PCM<br>(N = 724) | Control<br>(N = 3,620) | SMD | PCM<br>(N = 196) | Control<br>(N = 980) | SMD |
| **Age, mean years** | 68.3 | 68.6 | 0.026 | 69.5 | 69.3 | 0.021 |
| **Male, %** | 37.8% | 38.6% | 0.015 | 44.9% | 43.3% | 0.033 |
| **Low-income subsidy status, %** | 43.1% | 41.8% | 0.027 | 39.8% | 38.1% | 0.036 |
| **Charlson comorbidity index score, mean** | 3.6 | 3.6 | 0.016 | 4.4 | 4.5 | 0.037 |
| **Baseline medication use** |  |  |  |  |  |  |
| 90-day supply of maintenance medications, % | 97.5% | 97.5% | 0.002 | 96.4% | 96.6% | 0.011 |
| Total maintenance medications, mean | 10.9 | 10.8 | 0.007 | 11.8 | 11.9 | 0.032 |
| **Baseline resource use** |  |  |  |  |  |  |
| Emergency department visit, mean per 1000 members | 83.7 | 80.3 | 0.022 | 112.2 | 98.6 | 0.075 |
| Inpatient stays, mean per 1000 members | 41.7 | 42.8 | 0.014 | 77.0 | 79.0 | 0.019 |
| Total inpatient days, mean | 4.3 | 4.6 | 0.027 | 9.2 | 9.5 | 0.000 |
| **High VFA score, %** | 27.1% | 27.1% | 0.000 |  |  |  |
| **Completed CMR, %** | 32.0% | 32.0% | 0.000 | 42.9% | 42.9% | 0.000 |

PCM = pharmacy care management; SMD = standardized mean difference; CMR = comprehensive medical review Baseline characteristics were assessed during the 12-month period prior to the index date for PCM and Control members

The overall PCM cohort had a mean age of 68 years and 38% were male. Just under half of members received a low-income subsidy, and the average Charlson Comorbidity Index score was 3.6. Members were taking an average of 11 maintenance medications, and 98% had filled at least one 90-day supply in the baseline period; 32% of PCM members had completed a CMR in the baseline period.

The high VFA subgroup comprised 27% of PCM members and their matched controls. Compared to all PCM members, the high VFA group was slightly older, was more likely to be male, had a slightly lower proportion with low-income subsidy, had a higher comorbidity score, took more medications at baseline, and were less likely to have received a 90-day supply; they also were more likely to have completed a CMR in the baseline period. High VFA members also experienced substantially higher rates of ED visits and inpatient stays in the baseline period.

### Cost of Care

The impact of the PCM program on pharmacy, medical, and overall health spending is summarized in **Table 2**. Among all PCM members, there was an increase in pharmacy spending compared to controls (absolute difference $50 PEPM; 95% CI: $15, $86) and an offsetting decrease in medical spending (absolute difference $158 PEPM; 95% CI: -$265, - $51). Overall, PCM members had $108 PEPM (95% CI: -$221, $5) lower total cost of care compared to controls after 12 months.

**Table 2.** Change in cost of care PEPM from baseline to 12 months after intervention.

| **Cost Category**<br>**(PEPM)** | **Absolute**<br>**Difference** | **95% CI** | ***P-value*** |
| --- | --- | --- | --- |
| **All Members** |  |  |  |
| Pharmacy costs | \$50 | (\$15, \$86) | 0.0046 |
| Medical costs | -\$158 | (-\$265, -\$51) | 0.0040 |
| Total | -\$108 | (-\$221, \$5) | 0.0618 |
| **High VFA Members** |  |  |  |
| Pharmacy costs | \$52 | (-\$19, \$130) | 0.1869 |
| Medical costs | -\$458 | (-\$678, -\$233) | <0.0001 |
| Total | -\$406 | (-\$645, -\$161) | 0.0003 |

PCM = pharmacy care management; PEPM = per enrollee per month; CI = confidence interval

Savings were greater in the high VFA subgroup **(Table 2)**. High VFA PCM members incurred $52 PEPM (95% CI: -$19, $130) more pharmacy spending but $458 PEPM (95% CI: -$678, - $233) less medical spending than controls, for a net decrease of $406 PEPM (95% CI: -$645, -$161) in total cost of care after 12 months.

### Health Services Use

PCM members experienced reductions in inpatient stays, while use of these health services was consistent over the study period for controls **(Table 3)**. There were no changes in the use of emergency department or urgent care visits. Over the 12-month follow-up period, PCM members experienced 15% fewer inpatient stays (p=0.008) per 1000 member months compared to control members. Among the high VFA group, members had 32% fewer inpatient stays (p<0.001); all resource use (inpatient stays, ED visits, and urgent care visits) was 17% lower among the high VFA group (p=0.016).

**Table 3.** Change in resource use from baseline to 12 months after intervention

| Resource Use Category<br>(monthly mean per 1000<br>members) | PCM |  | Control |  | Absolute<br>Difference | *P-value* | Relative<br>Difference | *P-value* |
| --- | --- | --- | --- | --- | --- | --- | --- | --- |
|  | Pre | Post | Pre | Post |  |  |  |  |
| **All Members** | **N = 724** | **N = 724** | **N = 3,620** | **N = 3,620** |  |  |  |  |
| Inpatient stays | 42 | 33 | 43 | 40 | -6 | 0.0112 | -15% | 0.0082 |
| Emergency dept. visits | 84 | 73 | 80 | 71 | -1 | 0.9053 | -1% | 0.9049 |
| Urgent care visits | 13 | 13 | 14 | 15 | -1 | 0.6369 | -5% | 0.6230 |
| All | 139 | 119 | 137 | 125 | -7 | 0.2117 | -6% | 0.2022 |
| **High VFA Members** | **N = 196** | **N = 196** | **N = 980** | **N = 980** |  |  |  |  |
| Inpatient stays | 77 | 39 | 79 | 59 | -18 | 0.0001 | -32% | 0.0001 |
| Emergency dept. visits | 112 | 90 | 99 | 87 | -11 | 0.3164 | -11% | 0.2898 |
| Urgent care visits | 14 | 13 | 13 | 11 | 0 | 0.9056 | 3% | 0.9062 |
| All | 203 | 142 | 190 | 158 | -29 | 0.0228 | -17% | 0.0156 |

PCM = pharmacy care management

### Medication Adherence

Medication adherence for PCM and control members is reported in **Table 4**. Use of PCM was associated with statistically significant increases in the percent of members adherent to renin-angiotensin system antagonist, statin, and oral antidiabetic therapy, with increases ranging from 7.3 to 12.9 percentage points (absolute) among all PCM members and 12.2 to 14.3 percentage points (absolute) for high VFA members.

**Table 4.** Change in absolute percent of members adherent (PDC≥80%) from baseline to 12 months after intervention.

| Drug Category<br>(% of members) | PCM | Control | Absolute<br>Difference | *P-value* |
| --- | --- | --- | --- | --- |
| **All Members** | **N = 724** | **N = 3,620** |  |  |
| Diabetes | 6.3% | -1.0% | 7.3% | 0.0009 |
| Renin-angiotensin system antagonists | 0.7% | -8.7% | 9.4% | <0.0001 |
| Statins | 11.6% | -1.4% | 12.9% | <0.0001 |
| **High VFA Members** | **N = 196** | **N = 980** |  |  |
| Diabetes | 10.7% | -1.6% | 12.2% | 0.0038 |
| Renin-angiotensin system antagonists | 3.5% | -10.9% | 14.3% | 0.0014 |
| Statins | 14.4% | 1.8% | 12.6% | <0.0001 |

PCM = pharmacy care management
PCM and Control columns represent the mean difference between the 12 months prior to index and the 12-month follow-up period
All Members (PCM): Diabetes (N=284); Renin-angiotensin system antagonists (N=412); Statins (N=476)
All Members (Control): Diabetes (N=1,427); Renin-angiotensin system antagonists (N=2,141); Statins (N=2,428)
High VFA Members (PCM): Diabetes (N=75); Renin-angiotensin system antagonists (N=115); Statins (N=125)
High VFA Members (Control): Diabetes (N=387); Renin-angiotensin system antagonists (N=598); Statins (N=649)

## DISCUSSION

This study evaluated a novel intervention for managing the health of medically complex and vulnerable MA members. Designed to augment more traditional population health management programs in MA, ours focused on medication management as a means to avoiding costly medical complications. Our intervention integrated multiple pharmacy care management services into a single program for eligible members and delivered it on an ongoing basis to targeted members who chose to enroll in the service. Among members who enrolled and used the PCM program for 12 months, we found the program created meaningful savings in medical services spending, in part due to a significant reduction in hospitalizations. This was in spite of an increase in average medication spending, which was a result of significantly improved adherence and the program’s monthly refill cadence.

The PCM program initially was offered to MA members taking 8+ medications, but we also assessed the relative value of targeting the PCM program based on the VFA score, which predicts potentially avoidable future health costs associated with medication adherence for each member. Results suggest that the PCM’s favorable effects on health spending, health services use, and medication adherence were amplified in the high VFA subgroup, and targeting of the program on the basis of this score in addition to number of medications yields greater savings on a per-enrolled member basis.

This program was not done in isolation; our results were incremental to the effects of standard, ongoing care management efforts employed by UPMC MA plans to improve quality ratings and lower costs of care, including CMRs, adherence campaigns, clinical programs for chronic conditions, accountable care incentives for providers, and case management for medically complex members. Against this backdrop, the results suggest a comprehensive and integrated pharmacy service tailored to the needs of medically complex and vulnerable MA members created additional value.

A number of other studies have suggested that improving the quality of medication use in medically complex and socially vulnerable populations may hold promise for reducing complications of chronic diseases.^23-29^ However, a recent Cochrane review^6^ of interventions aimed at improving the appropriate use of polypharmacy in older people found that reductions in inappropriate prescribing were not consistently linked to improvements in hospital admissions, medication-related problems, or health spending. Another Cochrane review of 50 studies of interventions to improve medication adherence in older community-dwelling adults taking multiple long-term medications^7^ found that variations in individual interventions studied, methodological quality, and reported results made it difficult to find strong evidence in support of any single intervention.

Our study builds on this previous research in important ways. For example, past studies suggest that any single intervention is unlikely to affect meaningful change in long-term prescribing and adherence behavior among medically complex and vulnerable seniors. We therefore used a sustained, multi-pronged program that combines dispensing and packaging services with clinical pharmacy intervention in members predicted to benefit most. Compared to many one-time or temporary interventions evaluated previously, our sustained PCM program yielded stronger effects on medication adherence, health services utilization, and medical costs.

Our results compare favorably to a recent evaluation of another long-term pharmacy service that provides adherence packaging for polypharmacy adults.^10^ In that study, the authors reported total cost savings in patients with at least 12 months of participation of $196 PEPM. Results in that study were not adjusted for patients’ enrollment in other pharmacy care management programs. By comparison, our study found greater increases in medication adherence, a significant reduction in inpatient admissions, and greater savings at 12 months in the high VFA subgroup.

The results of our analysis should be interpreted in light of some limitations. Our program was deployed in a medically complex MA population; results could differ in settings with less medically complex or younger members. Despite matching the PCM and control groups on characteristics known to affect medical resource utilization and spending, it is possible that unmeasured differences between the groups could explain these findings. For example, while we have attempted to distinguish the impact of the PCM program from other services that may have been offered concurrently (like disease management) by subjecting PCM and control members to the same eligibility criteria and matching on healthcare utilization, disease severity, and history of some care management programs, it is possible there were unmeasured interventions given preferentially to the same members receiving the PCM program.

In conclusion, we designed, implemented, and evaluated the impact of a novel pharmacy care management program for medically complex and vulnerable members of a MA plan. We found that compared with closely matched controls, the intervention improved adherence to chronic medications, reduced hospital admissions, and reduced medical spending in members receiving the program for 12 months. Pharmacy spend was higher for PCM members, attributable to the more frequent rate of prescription fills and higher rates of adherence, but was more than offset by reductions in medical costs. The PCM program’s overall cost savings were driven by larger reductions among members with high predicted cost savings opportunities (the high VFA subgroup), who had $406 PEPM lower total spending than their matched controls. This finding underscores the value of targeting the PCM program based on predicted savings potential, rather than merely the number of medications a member is taking.

## Data Availability

All data produced in the present work are contained in the manuscript

## EXHIBITS

## APPENDIX

**Figure e1.**
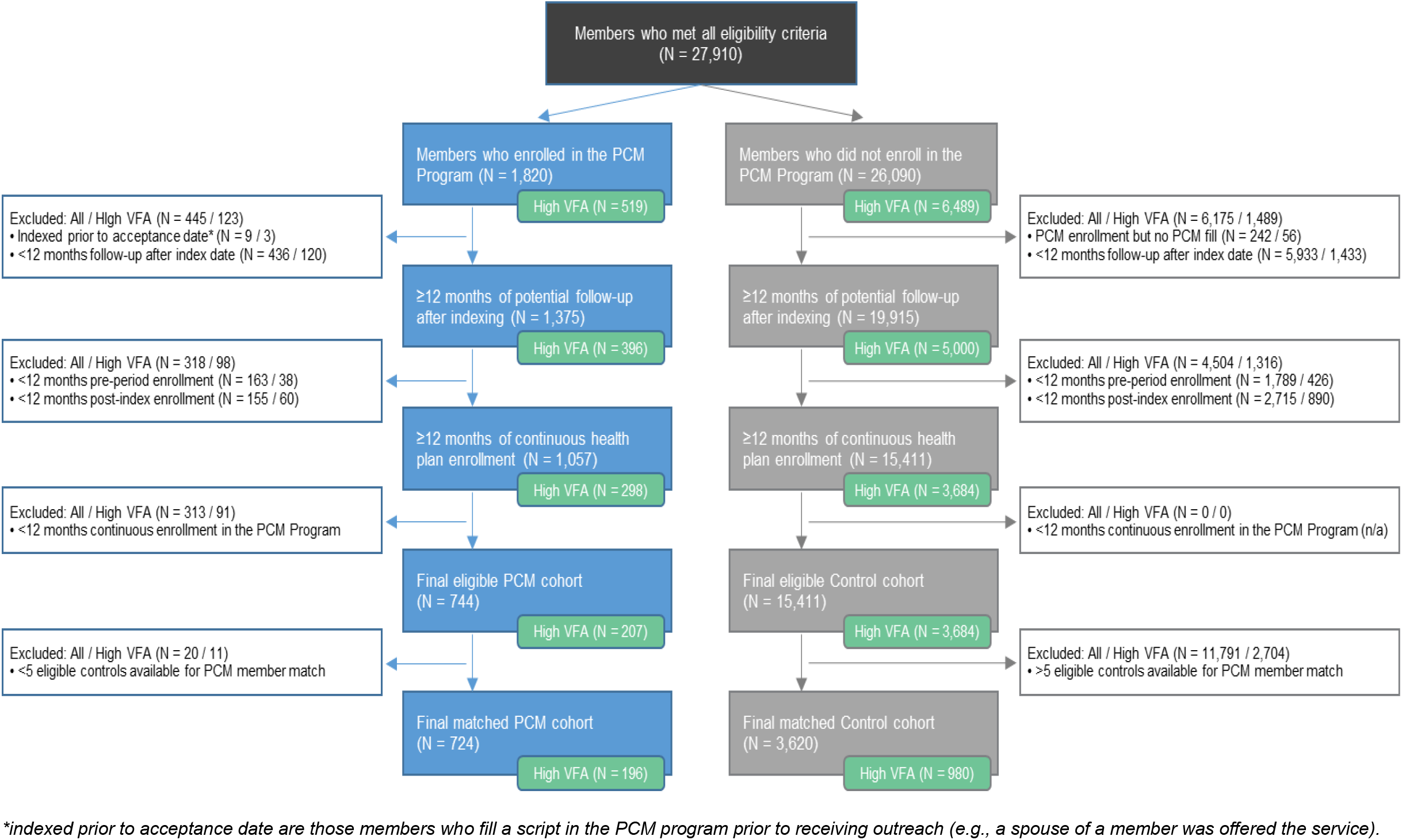
Attrition of members in the study cohorts.

**Table e2.** Baseline characteristics of Pharmacy Care Management members and potential controls, prior to matching.

| Baseline Characteristics | All Members |  |  | High VFA Members |  |  |
| --- | --- | --- | --- | --- | --- | --- |
|  | PCM<br>(N = 744) | Control<br>(N = 15,411) | SMD | PCM<br>(N = 207) | Control<br>(N = 3,684) | SMD |
| **Age, mean years** | 68.1 | 71.4 | 0.300* | 69.2 | 72.0 | 0.266* |
| **Male, %** | 38.0% | 41.9% | 0.079 | 44.9% | 46.6% | 0.034 |
| **Low-income subsidy status, %** | 43.8% | 35.2% | 0.177* | 41.1% | 35.2% | 0.121* |
| **Charlson comorbidity index score, mean** | 3.6 | 3.7 | 0.013 | 4.5 | 4.5 | 0.005 |
| **Baseline medication use** |  |  |  |  |  |  |
| 90-day supply of maintenance medications, % | 97.3% | 90.4% | 0.291* | 95.7% | 90.7% | 0.198* |
| Total maintenance medications, mean | 10.9 | 10.2 | 0.249* | 12.0 | 11.2 | 0.234* |
| **Baseline resource use** |  |  |  |  |  |  |
| Emergency department visit, mean per 1000 members | 85.7 | 68.9 | 0.112* | 115.5 | 94.3 | 0.114* |
| Inpatient stays, mean per 1000 members | 43.7 | 46.9 | 0.038 | 79.7 | 83.0 | 0.028 |
| Total inpatient days, mean | 4.5 | 5.7 | 0.084 | 9.6 | 11.1 | 0.070 |
| **High VFA score, %** | 27.8% | 23.9% | 0.090 |  |  |  |
| **Completed CMR, %** | 32.9% | 26.2% | 0.148* | 44.0% | 31.2% | 0.266* |

PCM = pharmacy care management; SMD = standardized mean difference; CMR = comprehensive medical review
Baseline characteristics were assessed during the 12-month period prior to index date for PCM and Control members.
\*SMD values >10%

